# Excessive Internet Use among Finnish Adolescents – Prevalence in 2017–2023 and Associations with Well-being

**DOI:** 10.64898/2026.10.06.26364845

**Authors:** Olli Kiviruusu, Silja Kosola, E. Juulia Paavonen

## Abstract

**Objective:** This study aimed at analyzing changes in the prevalence of excessive internet use and to analyze its associations with mental and cognitive health problems among adolescents.

**Method:** We used repeated cross-sectional data from the School Health Promotion study, a population-based nationwide classroom survey in Finland. All adolescents in grades 8–11 were invited, with a total of 602 226 responses in 2017–2023. Excessive internet use was measured with the five-item Excessive Internet Use Scale covering core behavioral addiction symptoms. Depressive symptoms, generalized anxiety, and social anxiety were measured with validated questionnaires. Problems in cognitive functioning were assessed with one-item indicators. Analyses were done with Chi-square tests and logistic regression.

**Results:** From 2017 to 2023, the prevalence of excessive internet use increased, especially among girls. The increasing trend was significantly (p<0.001) steeper among girls, with the OR for time (in years) being 1.12 in grades 8–9 and 1.14 in grades 10–11, while among boys the corresponding ORs were 1.00 and 1.04, respectively. Excessive internet use associated with short sleep, depressive, generalized anxiety, and social anxiety symptoms (ORs between 1.73–2.42 in boys; 1.92–2.26 in girls; p<0.001) and with difficulties in tasks requiring reading, writing, and mathematics, and difficulties in concentrating (ORs 1.84–2.31 in boys; 1.83–1.98 in girls; p<0.001).

**Conclusion:** The prevalence of excessive internet use showed an increasing trend, especially among girls. The associations of excessive internet use with mental health symptoms and cognitive challenges were clear.

## Introduction

In today’s digital age, the internet has become an integral part of adolescent life, shaping how young people communicate, learn, and entertain themselves. With social media platforms, video streaming services, and online gaming readily accessible, adolescents spend hours each day engaged in online activities.^1,2^ While the internet offers countless educational and social benefits, its extensive use among adolescents has raised concerns especially as to whether extensive use associates with their mental health, educational performance, and social development.^3–5^

For many adolescents, internet use remains a neutral or positive asset despite time spent online. Online life may become problematic due to inappropriate content, unsolicited contact or improper conduct (e.g. cyberbullying), or when internet use is associated with addiction-like behavior.^6,7^ The reported prevalence of problematic or excessive internet use (EIU) among adolescents varies greatly according to internet accessibility, population characteristics, sample representativeness, and definition used.^8–12^ For example, Nogueira-Lopez et al. reported a rate of 33% of EIU among Spanish adolescents when using a cutoff-based definition on a scale score (range 0–44), whereas the prevalence decreased to 3% when the same scale items were coded as dichotomous symptoms according to diagnostic criteria.^12^ In addition to variations in criteria and assessment tools, current understanding is further hindered by the relative scarcity of studies examining recent trends in EIU prevalence. Changes in EIU prevalence have mostly been reported before and during the COVID-19 pandemic,^13,14^ and studies extending to post-pandemic years are urgently needed.

During the past decade, both adolescent mental health and educational performance have declined significantly in most developed countries.^15–17^ In the United States, the prevalence of clinically diagnosed anxiety among adolescents increased from 10.0 % to 16.1 % between years 2016 and 2023,^16^ and significant increases in depressive symptoms have been observed since 2012.^18^ Similar trends in both mental health diagnoses and symptoms have been found in many high-income countries.^17^ Concurrently, according to the Programme for International Student Assessments (PISA), students in both the United States and Europe performed significantly worse in 2022 than before in both mathematics and reading.^15^ For example, mean performance had fallen by ten points in reading and nearly 15 points in mathematics since 2018—equivalent to about three-quarters of a year’s learning. The decline was more pronounced after the COVID-19 pandemic, although in many countries the downward trend had already begun before 2018.^15^ The rise in EIU is one proposed explanation for the observed declines in mental and cognitive health, as well as educational performance.^19^ This explanation has also been questioned and it is likely that the causal pathways are bidirectional and underlying mechanisms complex.^20^ Meta-analyses have indicated that EIU is associated with mental health^21^ and cognitive health.^22^ Nevertheless, to date, we found no population-based studies reporting on the associations of EIU with both mental and cognitive health outcomes, such as anxiety and the ability to concentrate.

The first aim of this population-based study was to report changes in EIU prevalence from 2017 to 2023 among Finnish youth. This also addresses the important question of whether the increases in the prevalence of EIU reported during the pandemic years were temporary, or whether the upward trend is continuing. In line with our previous report^13^ we hypothesize that the upward trend has continued. The second aim of our paper was to address the associations between EIU and cognitive and mental health outcomes. It is important to provide timely results on these associations. We hypothesized that adolescents with EIU would report poorer mental and cognitive health than their peers without EIU. We also address gender differences in these questions, since there are differences in the prevalence rates and their recent developments of both mental health problems as well as EIU.^13,23,24^ We hypothesize that the increasing trend of EIU is stronger among girls compared with boys.^13^

## Methods

### Design, Study Setting, and Sample

This population-based, repeated cross-sectional study utilized data from the School Health Promotion (SHP) study, a nationwide biennial survey managed by the Finnish Institute for Health and Welfare.^25^ The survey is based on total sampling – all schools in Finland and their students in grades 4, 5, and 8 to 11 are invited to participate. Students complete the online survey anonymously in classrooms under teacher supervision. Responding to the survey is voluntary and based on informed consent. For students under the age of 15, guardians are informed in advance, and they may decline their child’s participation. In this study, we utilized responses from 2017, 2019, 2021, and 2023, as the measure of excessive internet use (see below) has been included in the survey only since 2017. We also used responses only from grades 8–11, because the questionnaires for 4^th^ and 5^th^ graders did not include the outcome measures relevant for addressing the study questions.

Between 2017–2023, the number of participants ranged from 73 680 to 91 560 (coverage rates between years 63–75%) in grades 8 and 9, from 34 602 to 47 383 in academic high schools (coverage rates 54–71%), and from 19 215 to 26 625 (coverage rates 28–32%) in vocational education (more detailed account of number of participants and coverage rates for each survey year and school level is given in^25^). The number of participants in grades 8 to 11 per survey round ranged from 134 907 (in 2017) to 160 796 (in 2021), with a total of 602 226 responses in 2017–2023.

In Finland, education is compulsory, and tuition is free of charge from the age of seven to age 18 years. In grades 8 and 9, students are typically 14–16 years old. After grade 9, students apply to either academic high school or vocational education. In grades 10 and 11, students are usually 16–18 years old.

The institutional review board of the Finnish Institute for Health and Welfare has approved the study plan. We followed the Strengthening the Reporting of Observational Studies in Epidemiology (STROBE) guideline.

### Measures

All measures, except school level (grades 8–9, academic high school, or vocational education), were self-reported by the students.

#### Sociodemographic background

Students reported their gender as recorded in official documents (binary: boy or girl), age, grade (8 or 9; 10 or 11), and whether they lived in a household with both parents (yes/no). Family origin was based on questions about the students’ own and their parents’ countries of birth. A two-category variable for family origin was formed: 1) Finnish origin: the respondent and at least one parent born in Finland; and 2) Foreign origin/immigrant: both parents born abroad, and respondent born either in Finland or abroad.

Participants reported their parents’ level of education using options: “comprehensive school or equivalent” (meaning 9 years of education), “high school or vocational education” (12 years of education), “occupational studies in addition to high school or vocational education”, and “university, university of applied sciences or other higher education”. A “High parental education” variable (yes/no) was formed indicating that at least one parent had a university or higher education degree. Family financial situation was assessed using the question “How would you rate your family’s financial situation?” For statistical analyses, responses on a five-point scale were recoded into three categories: “good” (very/fairly good); “moderate” (moderate); and “poor” (fairly/very poor).

#### Excessive internet use

Excessive internet use (EIU) was assessed using the five-item Excessive Internet Use Scale (EIUS), a widely used measure in the European context.^26^ The five items cover components of behavioral addictions, namely cognitive and behavioral salience, tolerance, withdrawal symptoms, conflicts with other significant activities and social relationships, and relapse during attempts to regulate the behavior.^27^ The statements were: “I have tried unsuccessfully to spend less time online” (relapse), “I have spent less time than I should have with family, friends, or doing homework because of the time I spent online” (conflict), “I have caught myself surfing when I am not really interested” (tolerance), “I have felt bothered when I cannot be online” (withdrawal symptoms), and “I have gone without eating or sleeping because of the internet” (salience). Respondents were asked to evaluate how often they experienced each of the above on a four-point Likert scale: “never”, “not very often”, “fairly often”, and “very often”. The responses ‘fairly often’ and ‘often’ were interpreted as indicating the presence of a symptom, and the criteria for EIU required endorsement of the conflict symptom and at least three of the other symptoms, following procedures described in previous studies.^28^ Percentages of each item above the threshold meeting criteria for overall EIU are given in Supplementary Table S2.

#### Mental health outcomes

Sleep duration was derived from two questions: “At what time do you usually go to bed?” and “At what time do you usually wake up?” Response options for bedtime and wake-up times were provided at half-hour intervals. A dichotomous variable for sleeping less than seven hours per weeknight (yes/no) was calculated.

Depressive symptoms were measured with the Patient Health Questionnaire 2 (PHQ-2), which enquires about the frequency of depressed mood and anhedonia over the last two weeks.^29^ Response options range from “not at all” to “nearly every day” on a four-point Likert scale. Total scores range from 0 to 6, and a cut-off ≥3 was used as suggested among adolescents to be optimal for detecting depression.^30^

The seven-item Generalized Anxiety Disorder scale (GAD-7) was used to measure symptoms of generalized anxiety.^31^ Items are scored on a 4-point Likert scale from “not at all” to nearly every day”. Total scores range from 0 to 21, and a cut-off of ≥10 was used to indicate moderate to severe generalized anxiety symptom, as supported by validation studies in both adolescent and adult populations.^31,32^

Social anxiety was assessed using the three-item Mini-Social Phobia Inventory (Mini-SPIN), where adolescents evaluate the severity of avoidance behavior and fear of embarrassment relating to social situations during the past week.^33^ All items are scored on a five-point Likert scale from “not at all” to “extremely”, and total scores range from 0 to 12. A cut-off score ≥6 has been validated among Finnish adolescents to detect social anxiety disorder.^34^

#### Cognitive health outcomes

Difficulties in concentration, reading, writing, and mathematics were assessed through single items. For concentration, the item read: “Do you have difficulties concentrating even when the task is fun?” with response options “no difficulties”, “some difficulty”, “a lot of difficulty”, and “cannot do at all”. For reading, writing, and mathematics the item enquired about difficulties in tasks requiring these skills and the response options were “not at all”, “fairly little”, “quite a lot”, and “very much”. For all four measures the two lowest and two highest responses were respectively combined for a dichotomous variable.

### Statistical Analysis

Analyses were done using IBM SPSS Statistics 29.0 software and reported separately for boys, girls, and the total sample.

We excluded 5647 (0.9%) cases between 2017 and 2023 due to implausible or unreliable responding, namely reporting 1) in three questions on disabilities that they were “completely unable” to see, hear, and walk, or 2) that during the school year they had experienced two or more accidents requiring a visit to the school nurse or a doctor in each of the following four contexts: during recess, physical education lessons, other classes, and on the way to school (i.e., at least eight accidents in total). In the SHP study, addressing implausible or unreliable responding is standard practice, although the specific methods used vary somewhat depending on the studied survey years and subgroups.^25^ The variables used in the present study were available for all included survey years. After these exclusions, 596 441 participants in 2017–2023 were included in the analyses.

For the prevalence of EIU, percentages with 95% confidence intervals (CI) were reported for two groups (grades 8–9, and 10–11). Logistic regression was used to analyze the effect of time (linear, in years) on the prevalence of EIU. Unadjusted models were subsequently adjusted for sociodemographic background factors. Gender differences in the effects were analyzed using time × gender interaction terms.

Associations between EIU and the outcomes in 2023 were first analyzed by cross-tabulating the outcomes against EIU status (with or without), and group differences were tested using the Chi-square test. The associations were further analyzed using logistic regression adjusting for sociodemographic background factors. In the logistic regression models EIU was the independent variable, and mental and cognitive outcomes were the dependent variables. Gender differences in the effects were analyzed using EIU × gender interaction terms.

The amount of missing information was relatively small (Table 1). Results are presented from analyses with all available data for a given analysis. Sensitivity analyses were done using complete case data, meaning that only cases with no missing values in the variables relevant to the study question were included.

**Table 1.**
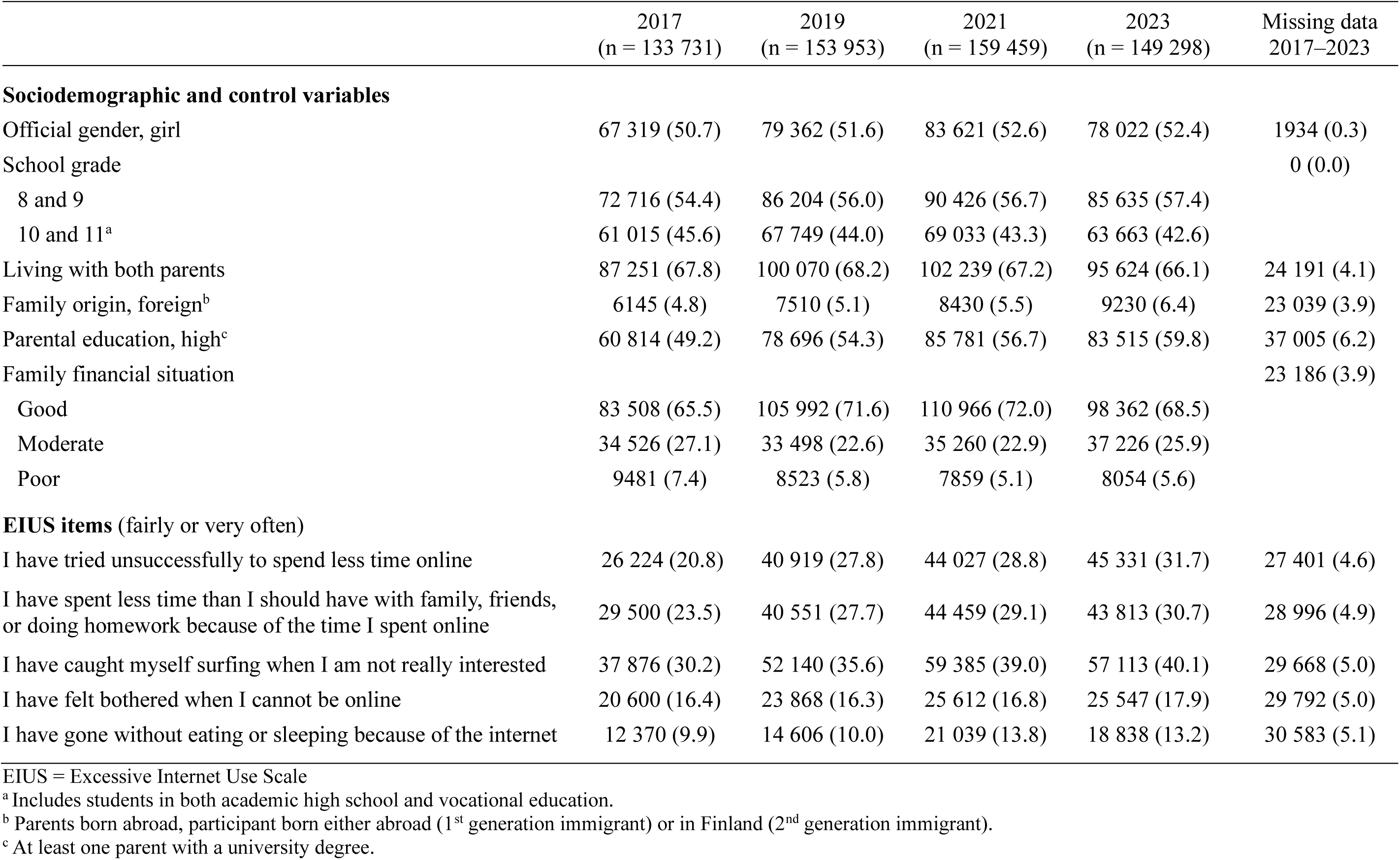
Study participants and descriptive statistics by study year, n (%).

## Results

Sociodemographic characteristics of the study sample and prevalence of EIU symptoms by study year are presented in Table 1. The prevalence of all EIU symptoms increased from 2017 to 2023. Symptoms relating to surfing even when not interested (40.1% in 2023) and unsuccessful efforts to limit time online (31.7% in 2023) were most frequently reported. (For distributions of the EIUS items, see Table S1 in the Supplement).

Among girls, EIU increased steadily across all timepoints (Figure 1 and Table S3 in the Supplement). Among girls in grades 8–9, EIU increased from 7.7% (95% CI 7.5–8.0%) in 2017 to 14.0% (95% CI 13.7–14.4%) in 2023, highest of all study groups. Among girls in grades 10–11, the prevalence of EIU more than doubled and the effect of time on EIU (OR 1.14; 95% CI 1.13–1.15) was strongest in this group (Table 2). Among boys, changes in EIU were modest, and the effect of time on EIU significantly smaller than among girls as indicated by moderation analyses by gender.

**Figure 1.**
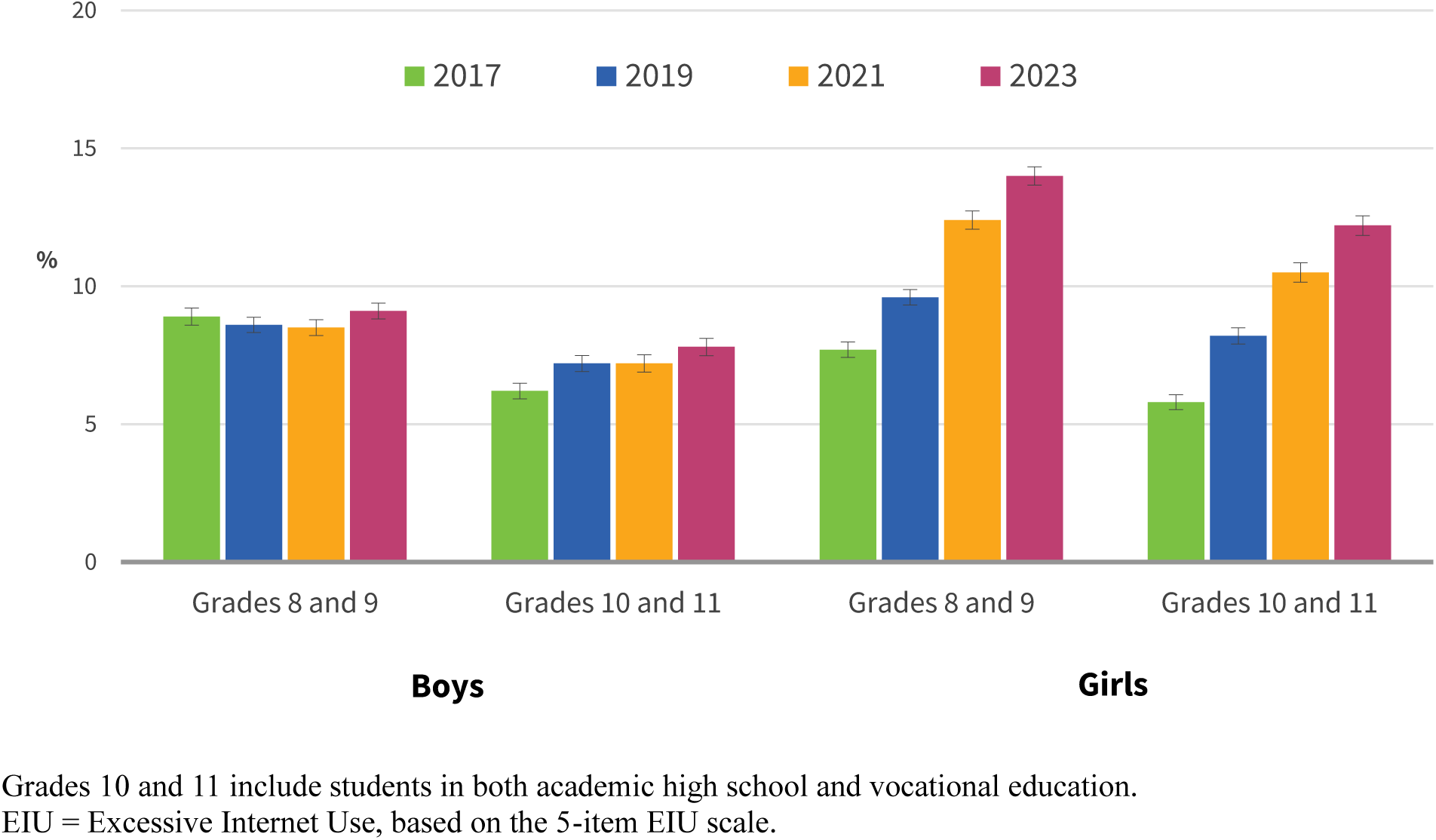
Prevalence and 95% confidence intervals of EIU between 2017 and 2023 by gender and grade.

**Table 2.** Effect of linear time (years) on EIU between 2017 and 2023 by gender and grade. Odds ratios (OR) from logistic regression models.

| Subgroup | Unadjusted model |  | Adjusted model<sup>b</sup> |  |
| --- | --- | --- | --- | --- |
|  | OR (95% CI) | p | OR (95% CI) | p |
| **Boys** |  |  |  |  |
| Grades 8 and 9 | 1.00 (1.00–1.01) | 0.419 | 1.01 (1.00–1.01) | 0.271 |
| Grades 10 and 11<sup>a</sup> | 1.04 (1.03–1.05) | < 0.001 | 1.04 (1.03–1.05) | < 0.001 |
| **Girls** |  |  |  |  |
| Grades 8 and 9 | 1.12 (1.11–1.13)<sup>c</sup> | < 0.001 | 1.12 (1.11–1.13)<sup>c</sup> | < 0.001 |
| Grades 10 and 11<sup>a</sup> | 1.14 (1.13–1.15)<sup>c</sup> | < 0.001 | 1.14 (1.13–1.15)<sup>c</sup> | < 0.001 |
| **Total** |  |  |  |  |
| Grades 8 and 9 | 1.07 (1.06–1.07) | < 0.001 | 1.07 (1.07–1.08) | < 0.001 |
| Grades 10 and 11<sup>a</sup> | 1.10 (1.09–1.10) | < 0.001 | 1.10 (1.09–1.11) | < 0.001 |

<sup>a</sup> Includes students in both academic high school and vocational education.
<sup>b</sup> Analyses were adjusted for grade (8, 9, 10, and 11), living with both parents, family origin, parental education, and family financial situation, in total sample models also for gender.
<sup>c</sup> Moderation by gender: effect significantly different from the corresponding effect in boys, $p < 0.001$
EIU = Excessive Internet Use, based on the 5-item EIU scale.

EIU was associated with short sleep duration, depressive symptoms, generalized and social anxiety symptoms, and difficulties in reading, writing, mathematics, and concentration among both boys and girls (Table 3). The associations remained significant when the results were adjusted for sociodemographic background factors (Table 4). The associations of EIU with short sleep and depressive symptoms were stronger among girls, while the association with generalized anxiety symptoms was slightly stronger among boys as indicated by statistically significant EIU × gender interaction terms (Table 4).

**Table 3.** Associations of EIU with short sleep, and mental and cognitive health symptoms among students in grades 8–11 in 2023.

| Outcome | Boys |  |  | Girls |  |  | Total |  |  |
| --- | --- | --- | --- | --- | --- | --- | --- | --- | --- |
|  | No EIU<br>(n = 60 956) | With EIU<br>(n = 5704) | p<sup>a</sup> | No EIU<br>(n = 65 810) | With EIU<br>(n = 10 011) | p<sup>a</sup> | No EIU<br>(n = 126 766) | With EIU<br>(n = 15 715) | p<sup>a</sup> |
|  | n (%) | n (%) |  | n (%) | n (%) |  | n (%) | n (%) |  |
| Sleep per weeknight <7 h | 6777 (11.2) | 1056 (19.1) | < 0.001 | 8769 (13.4) | 2488 (25.1) | < 0.001 | 15 546 (12.4) | 3544 (22.9) | < 0.001 |
| Depressive symptoms | 6051 (10.1) | 1036 (18.7) | < 0.001 | 18 603 (28.6) | 4757 (48.2) | < 0.001 | 24 654 (19.7) | 5793 (37.6) | < 0.001 |
| Generalized anxiety symptoms | 3951 (6.5) | 833 (14.8) | < 0.001 | 19 778 (30.1) | 4898 (49.1) | < 0.001 | 23 729 (18.8) | 5731 (36.7) | < 0.001 |
| Social anxiety symptoms | 11 610 (19.3) | 1582 (28.6) | < 0.001 | 31 500 (48.3) | 6353 (64.3) | < 0.001 | 43 110 (34.4) | 7935 (51.5) | < 0.001 |
| Difficulties in tasks requiring |  |  |  |  |  |  |  |  |  |
| Reading | 10 842 (17.9) | 1664 (29.4) | < 0.001 | 19 305 (29.4) | 4459 (44.8) | < 0.001 | 30 147 (23.9) | 6123 (39.2) | < 0.001 |
| Writing | 11 645 (19.2) | 1699 (30.1) | < 0.001 | 17 082 (26.0) | 4040 (40.6) | < 0.001 | 28 727 (22.7) | 5739 (36.8) | < 0.001 |
| Mathematics | 10 348 (17.1) | 1536 (27.2) | < 0.001 | 22 562 (34.4) | 5039 (50.7) | < 0.001 | 32 910 (26.1) | 6575 (42.1) | < 0.001 |
| Difficulties concentrating | 4619 (7.6) | 904 (16.1) | < 0.001 | 13 656 (20.8) | 3615 (36.4) | < 0.001 | 18 275 (14.5) | 4519 (29.1) | < 0.001 |

<sup>a</sup> Difference between groups with and without EIU tested with Chi-square test.
EIU = Excessive Internet Use, based on the 5-item EIU scale.

**Table 4.** Odds ratios (OR) from adjusted^a^ logistic regression models for EIU as a predictor of mental and cognitive health outcomes among students in grades 8–11 in 2023.

| Outcome (dependent variable) | Boys |  | Girls |  | Total |  |
| --- | --- | --- | --- | --- | --- | --- |
|  | OR (95% CI)<sup>b</sup> | p | OR (95% CI)<sup>b</sup> | p | OR (95% CI)<sup>b</sup> | p |
| Sleep per weeknight <7 h | 1.73 (1.59–1.88) | < 0.001 | 2.10 (1.99–2.21)<sup>c</sup> | < 0.001 | 1.98 (1.89–2.07) | < 0.001 |
| Depressive symptoms | 1.98 (1.82–2.15) | < 0.001 | 2.26 (2.16–2.37)<sup>d</sup> | < 0.001 | 2.20 (2.11–2.29) | < 0.001 |
| Generalized anxiety symptoms | 2.42 (2.21–2.65) | < 0.001 | 2.19 (2.09–2.29)<sup>e</sup> | < 0.001 | 2.24 (2.15–2.33) | < 0.001 |
| Social anxiety symptoms | 1.76 (1.64–1.88) | < 0.001 | 1.92 (1.83–2.01) | < 0.001 | 1.87 (1.80–1.94) | < 0.001 |
| Difficulties in tasks requiring |  |  |  |  |  |  |
| Reading | 1.94 (1.81–2.07) | < 0.001 | 1.93 (1.85–2.02) | < 0.001 | 1.93 (1.86–2.01) | < 0.001 |
| Writing | 1.87 (1.75–2.00) | < 0.001 | 1.93 (1.84–2.02) | < 0.001 | 1.90 (1.83–1.98) | < 0.001 |
| Mathematics | 1.84 (1.71–1.97) | < 0.001 | 1.97 (1.88–2.06) | < 0.001 | 1.93 (1.86–2.00) | < 0.001 |
| Difficulties concentrating | 2.31 (2.12–2.52) | < 0.001 | 2.13 (2.04–2.24) | < 0.001 | 2.18 (2.09–2.27) | < 0.001 |

<sup>a</sup> Adjusted for grade (8, 9, 10, and 11), living with both parents, family origin, parental education, and family financial situation, in total sample models also for gender.
<sup>b</sup> Separate model for each outcome; OR for EIU predicting the outcome.
<sup>c,d,e</sup> Moderation by gender: effect significantly different from the corresponding effect in boys: <sup>c</sup> $p < 0.001$ , <sup>d</sup> $p < 0.01$ , <sup>e</sup> $p < 0.05$ .
EIU = Excessive Internet Use, based on the 5-item EIU scale.

Sensitivity analyses using only complete cases replicated the main results (Table S4 and Table S5 in the Supplement).

## Discussion

In this population-based study, EIU increased steadily among adolescent girls from 2017 to 2023, while changes among boys were modest. EIU was associated with short sleep, symptoms of depression and anxiety, and cognitive difficulties at school among both boys and girls. All associations were highly significant, regardless of sociodemographic factors.

The prevalence of EIU increased over the six-year period among Finnish adolescents. The rising prevalence rates are in line with studies preceding COVID-19 ^35^ or extending to the pandemic years.^11,13^ Our findings show, and in line with our hypothesis, that EIU prevalences estimates have continued to increase in post-pandemic time. Contrary to some previous studies,^11^ the pandemic time did not seem to affect the trend of EIU among Finnish adolescents, as was already suggested in our earlier study.^13^ If anything, among boys between grades 8 and 11, there was a temporary standstill in the trend during the pandemic.

The increased prevalence of self-reported EIU among adolescent girls from 2017 to 2023 is remarkable. And as we hypothesized the increase among girls was significantly more rapid compared to boys. By 2023, adolescent girls reported higher prevalence of excessive internet use (EIU) than boys. This differs from earlier studies, which usually found higher rates among men.^35,36^

However, those studies were based on samples collected in 2017 or earlier, prior to the onset of our study period, and prior to the widespread adoption of social media platforms. The shift toward a female predominance in EIU among Finnish adolescents is plausible given the central role of social media in adolescents’ lives, particularly among girls.¹ Social media use has increased rapidly in recent years, and this increase has been stronger among girls than boys.^37^ In contrast, gaming remains more prevalent among boys than girls, and a recent meta-analysis did not find evidence of increasing trends in internet gaming disorder prevalence.^38^ Thus, the observed diverging trends in EIU prevalence between girls and boys may reflect gender differences in internet use profiles and recent developments in the intensity and patterns of use within these online environments.^37,39^

In line with our hypothesis, the associations between EIU and mental and cognitive health indicators were clear and consistent throughout the studied measures. These findings align well with previous studies regarding the associations of EIU with mental health problems ^21^ and cognitive difficulties.^22^ In comparison to a simple measure of screen time, EIU depicts the negative consequences which adolescents experience in relation to internet use. Thus, the clear associations might be due to the used construct – while screentime may be neutral or beneficial, EIU is by definition something negative. This suggests that measurement of more EIU-like constructs in addition to screentime may be useful.^39^

One of the most popular explanations for the harms of excessive internet and social media use is the sleep displacement theory.^40^ Sufficient sleep is vital for growth, mental health, and cognitive performance, and internet use prior to bedtime may delay sleep.^41^ Digital media use is related to shorter sleep duration and worse sleep quality.^42^ Also in the present study short sleep was more frequent among adolescents with EIU than their peers. However, it should be noted that reduced sleep is already partly captured by the salience item of the EIUS; therefore, the association between sleep and EIU also reflects this conceptual overlap between the measures.

Many other mechanisms may also explain the links between internet use and mental and cognitive health. Mental health may be affected by the content, contacts, conduct, and contracts adolescents face online,^6^ and psychiatric symptoms may spread through social contagion online.^43^ Cognitive health outcomes may be affected by media multitasking or the rapid tempo and addictive nature of online material.^44,45^ The power of social media algorithms is suggested especially in one of the EIU items: 32% of adolescents (mainly aged 14–18 years) had failed to reduce time spent online in 2023, up from 21% in 2017. This contradicts the previously proposed age of 13 years as suitable for social media use.^5^ In addition to the mechanisms discussed above, it is important to note that the relationship between internet use and mental and cognitive health is likely bidirectional. For instance, adolescents experiencing depression or anxiety may be more susceptible to developing EIU, in addition to — or rather than — EIU contributing to the onset of mental health symptoms.^46,47^

The subjective experience of EIU and associated mental and cognitive health symptoms may be reflected in health care service demand. This demand may in part explain the increase in diagnoses of attention-deficit/hyperactivity disorder, anxiety, depression, and even self-harm, which may all be secondary to EIU.^48,49^ Screening for EIU symptoms in clinical practice among adolescents presenting with mental or cognitive health symptoms may help target support and prevent more severe problems.

The rise in EIU and mental health symptoms as well as the decrease in academic achievement all began already before the COVID-19 pandemic.^4,8,13,15,18^ The pandemic further exacerbated these developments, and recovery has been slow.^24^ Our findings are in line with concerns that the worsening trends in adolescent mental and cognitive health may be linked to increased screen time and addiction-like symptoms, such as EIU. However, more rigorous study designs are needed to uncover potential causal pathways and the underlying mechanisms connecting EIU with mental and cognitive health in adolescents. These are critical questions that future research should aim to address.

### Strengths and Limitations

The strengths of our study include a large, nationally representative sample, population-based design, high response rates across timepoints, and the use of validated measures for assessing mental health symptoms. In addition to mental health symptoms, study participants also provided concurrent estimates of their cognitive health at school. Nonetheless, some limitations warrant consideration. Due to the use of repeated anonymous cross-sectional surveys, individual-level longitudinal changes could not be assessed. Despite the large sample size, the results may not represent all adolescents, such as adolescents absent from school or unwilling to participate. Thus, to the extent that nonparticipation is related to EIU, our prevalence estimates may be biased depending on the rate of nonparticipation. Student coverage in vocational institutions was notably lower than in other groups, likely due to data collection overlapping with their practical training periods, while possibly also due to lower student engagement. We had no data regarding the presence or absence of psychiatric disorders or learning disabilities. Using single-item measures for cognitive health is suboptimal, although the face validity of the items appears acceptable, as terms such as reading and writing are clear, simple, and therefore age-appropriate and easy for participants to understand. Our measure of sleep does not fully capture actual sleep duration, as it is based solely on reported bedtimes and wake times and does not account for interrupted sleep. The definition of EIU used in this study is quite conservative to avoid overestimating its prevalence. EIU represents the subjective experience of adolescents on whether their time spent online is associated with problems, but we have no measure of screentime or details on how adolescents spent their time online. Questionnaire based studies are prone to recall and social desirability bias, though anonymity and teacher supervision probably reduced the latter. In a study among adolescents, the correlation between self-reported and objectively measured screen time was 0.42,^1^ illustrating the challenges associated with the reliability of self-reported screen-time measures and related activities. Although such objective approaches are not feasible in large-scale population surveys, future studies could use smartphone technologies and Ecological Momentary Assessments (EMA) to obtain a more nuanced picture of adolescents’ online experiences and to better understand how these experiences may relate to the onset of behavioral addictions. Finally, the observed associations may be influenced by unmeasured variables, confounding or mediating factors, such as adolescent sleep quality, substance use or parental mental health and internet use.^50^ Causality or the direction of effects cannot be determined based on these data.

## Conclusions

In this study, the prevalence of EIU increased significantly from 2017 to 2023 among adolescent girls. EIU was associated with mental and cognitive health outcomes. The increased prevalence of EIU is of concern and warrants its constant monitoring among adolescents. Further studies are needed to evaluate risk factors beyond EIU and to evaluate effectiveness of intervention studies. Our findings call for prompt action to support adolescent health and prevent the negative impacts of digital life, which are increasingly reflected in the symptoms of EIU.

## Data Availability

Finnish Institute for Health and Welfare (THL) has collected the data and has the rights for the data. THL produces public statistical reports and interactive reports, but the data is confidential and thus not publicly available. Researchers can apply the data from Findata (https://findata.fi/en/) or the 2019 and 2025 data from Finnish Social Science Data Archive (https://www.fsd.tuni.fi/en/).

https://www.fsd.tuni.fi/en/

## Notes

### Competing Interest Statement

The authors have declared no competing interest.

### Author Declarations

The institutional review board of the Finnish Institute for Health and Welfare gave ethical approval for this work.

### Funding

- Juho Vainio Foundation: 202500482
- Foundation for Pediatric Research

## References

1. Kosola S, Mörö S, Holopainen E. Smartphone use and well-being of adolescent girls: A population-based study. Arch Dis Child. 2024;109(7):576–581.

2. Madigan S, Eirich R, Pador P, McArthur BA, Neville RD. Assessment of changes in child and adolescent screen time during the COVID-19 pandemic: A systematic review and meta-analysis. JAMA Pediatr. 2022;176(2):1188–1198.

3. Santos RMS, Mendes CG, Bressani GYS, et al. The associations between screen time and mental health in adolescents: A systematic review. BMC Psychol. 2023;11:127.

4. Qu G, Hu W, Meng J, et al. Associations between screen time and developmental and behavioral problems among children in the United States: Evidence from 2018 to 2020 NSCH. J Psychiatr Res. 2023;161:140–149.

5. Montag C, Demetrovics Z, Elhai JD, et al. Problematic social media use in childhood and adolescence. Addict Behav. 2024;153:107980.

6. Livingstone S, Stoilova M. The 4Cs: Classifying online risk to children. (CO:RE Short Report Series on Key Topics). Hamburg: Leibniz-Institut für Medienforschung | Hans-Bredow-Institut (HBI); CO:RE – Children Online: Research and Evidence. 2021. doi: 10.21241/ssoar.71817.

7. Nagata JM, Helmer CK, Al-Shoaibi AA. Beyond screen time – addictive screen use patterns and adolescent mental health. JAMA. 2025; online ahead of print doi: 10.1001/jama.2025.8135.

8. Boer M, van den Eijnden RJJM, Boniel-Nissim M, et al. Adolescents’ intense and problematic social media use and their well-being in 29 countries. J Adolesc Health. 2020;66(6):S89–S99.

9. Fineberg NA, Menchón JM, Hall N, et al. Advances in problematic usage of the internet research–A narrative review by experts from the European network for problematic usage of the internet. Compr Psychiatry. 2022;118:152346.

10. Joseph J, Varghese A, Vijay VR, et al. Problematic internet use among school-going adolescents in India: A systematic review and meta-analysis. Indian J Community Med. 2022;47(3):321–327.

11. Liu X, Gui Z, Chen Z, et al. Global prevalence of internet addiction among university students: a systematic review and meta-analysis. Curr Opin Psychiatry. 2025;38(3):182–199.

12. Noguiera-Lopez A, Rial-Boubeta A, Guadix-García I, Villanueva-Blasco VJ, Billieux J. Prevalence of problematic internet use and problematic gaming in Spanish adolescents. Psychiatry Res. 2023;326:115317.

13. Kiviruusu O. Excessive internet use among Finnish young people between 2017 and 2021 and the effect of COVID-19. Soc Psychiatry Psychiatr Epidemiol. 2024;59(12):2291–2301.

14. Kamasak T, Topbas M, Cansu A, et al. An investigation of changing attitudes and behaviors and problematic internet use in children aged 8 to 17 years during the COVID-19 pandemic. Clin Pediatr. 2022;61(2):194–205.

15. OECD. PISA 2022 results (volume I): The state of learning and equity in education. PISA, OECD Publishing, Paris 2023. doi: 10.1787/53f23881-en.

16. Sappenfield O, Alberto C, Minnaert J, Donney J, Lebrun-Harris L, Ghandour R. National Survey of Children’s Health. Adolescent Mental and Behavioral Health 2023. Health Resources and Services. Administration, Data brief October 2024. Available online at: https://mchb.hrsa.gov/sites/default/files/mchb/data-research/nsch-data-brief-adolescent-mental-behavioral-health-2023.pdf. Accessed July 8, 2025.

17. McGorry PD, Mei C, Dalal N, et al. The Lancet Psychiatry Commission on youth mental health. The Lancet Psychiatry. 2024;11(9):731–774.

18. Keyes KM, Gary D, O’Malley PM, HamiltonA, Schulenberg J. Recent increases in depressive symptoms among US adolescents: trends from 1991 to 2018. Soc Psychiatry Psychiatr Epidemiol. 2019;54:987–996.

19. Haidt J. The anxious generation: How the great rewiring of childhood is causing an epidemic of mental illness. New York, NY: Penguin Press; 2024.

20. Orben A. Teenagers, screens and social media: a narrative review of reviews and key studies. Soc Psychiatry Psychiatr Epidemiol. 2020;55(4):407–414.

21. Cai Z, Mao P, Wang Z, Wang D, He J, Fan X. Associations between problematic internet use and mental health outcomes of students: a meta-analytic review. Adolesc Res Rev. 2023;8(1):45–62.

22. Ioannidis K, Grant JE, Chamberlain SR. Problematic usage of the internet and cognition. Curr Opin Behav Sci. 2022;44:101104.

23. Su W, Han X, Yu H, Wu Y, Potenza MN. Do men become addicted to internet gaming and women to social media? A meta-analysis examining gender-related differences in specific internet addiction. Comput Human Behav. 2020;113:106480.

24. Kiviruusu O, Ranta K, Lindgren M, et al. Mental health after the COVID-19 pandemic among Finnish youth: a repeated, cross-sectional, population-based study. Lancet Psychiatry. 2024;11(6):451–460.

25. Helenius J, Kivimäki H. Well-being of children and young people – School Health Promotion study 2023. Statistical report 50/2023. Finnish Institute for Health and Welfare. https://urn.fi/URN:NBN:fi-fe20230913124233.

26. Skarupová K, Ólafsson K, Blinka L. Excessive internet use and its association with negative experiences: quasi-validation of a short scale in 25 European countries. Comput Hum Behav. 2015;53:118–123.

27. Griffiths M. A “components” model of addiction within a biopsychosocial framework. J Subst Use. 2005;10(4):191–197.

28. Lukavska K, Vacek J, Gabhelík R. The effects of parental control and warmth on problematic internet use in adolescents: A prospective cohort study. J Behav Addict. 2020;9(3):664–675.

29. Kroenke K, Spitzer RL, Williams JBW. The Patient Health Questionnaire-2: validity of a wo-item depression screener. Med Care. 2003;41(11):1284–1292.

30. Richardson LP, Rockhill C, Russo JE, et al. Evaluation of the PHQ-2 as a brief screen for detecting major depression among adolescents. Pediatrics. 2010;125(5):e1097–1103.

31. Spitzer RL, Kroenke K, Williams JBW, Löwe B. A brief measure for assessing generalized anxiety disorder: the GAD-7. Arch Intern Med. 2006;166(10):1092–1097.

32. Guzick A, Storch EA, Smárason O, et al. Psychometric properties of the GAD-7 and PROMIS-Anxiety-4a among youth with depression and suicidality: Results from the Texas youth depression and suicide research network. J Psychiatr Res. 2024;170:237–244.

33. Connor KM, Kobak DA, Churchill LE, Katzelnick D, Davidson JR. Mini-SPIN: a brief screening assessment for generalized social anxiety disorder. Depress Anxiety. 2001;14(2):137–140.

34. Ranta K, Kaltiala-Heino R, Rantanen P, Marttunen M. The Mini-Social Phobia Inventory: psychometric properties in an adolescent general population sample. Compr Psychiatry. 2012;53(5):630–637.

35. Shao YJ, Zheng T, Wang YQ, Liu L, Chen Y, Yao Y. Internet addiction detection rate among college students in the People’s Republic of China: a meta-analysis. Child Adolesc Psychiatry Ment Health. 2018;12:25.

36. Anderson EL, Steen E., Stavropoulos V. Internet use and problematic internet use: A systematic review of longitudinal research trends in adolescence and emergent adulthood. Int J Adolesc Youth. 2017;22(4):430–454.

37. Boniel-Nissim M, Marino C, Galeotti T, et al. A focus on adolescent social media use and gaming in Europe, central Asia and Canada. Health Behaviour in School-aged Children international report from the 2021/2022 survey. Volume 6. Copenhagen: WHO Regional Office for Europe; 2024.

38. Pan YC, Chiu YC, Lin YH. Systematic review and meta-analysis of epidemiology of internet addiction. Neurosci Biobehav Rev. 2020;118:612–622.

39. Rega V, Gioia F, Boursier V. Problematic media use among children up to the age of 10: a systematic literature review. Int J Environ Res Public Health. 2023;20(10):5854.

40. Ellithorpe ME, Eden A, Ulusoy E, Wirz D, Grady S. Is bedtime media use good or bad? A competitive analysis between the sleep displacement hypothesis and the media recovery hypothesis. Media Psychology. 2024;28(4):516–547.

41. Crowley SJ, Wolfson AR, Tarokh L, Carskadon MA. An update on adolescent sleep: New evidence informing the perfect storm model. J Adolesc. 2018;67:55–65.

42. Brautsch LA, Lund L, Andersen MM, Jennum PJ, Folker AP, Andersen S. Digital media use and sleep in late adolescence and young adulthood: A systematic review. Sleep Med Rev. 2023;68:101742.

43. Kramer ADI, Guillory JE, Hancock JT. Experimental evidence of massive-scale emotional contagion through social networks. PNAS. 2014;111(24):8788–8790.

44. Mo C-Y, Wang C, Dai J, Jin P. Video playback speed influence on learning effect from the perspective of personalized adaptive learning: a study based on cognitive load theory. Front Psychol. 2022;13:839982.

45. Firth J, Torous J, Stubbs B, et al. The “online brain”: how the internet may be changing our cognition. World Psychiatry 2019;18(2):119–129.

46. Kardefelt-Winther D. A conceptual and methodological critique of internet addiction research: Towards a model of compensatory internet use. Comput Hum Behav. 2014;31:351–354.

47. Odgers CL, Jensen MR. Annual research review: Adolescent mental health in the digital age: Facts, fears, and future directions. J Child Psychol Psychiatry. 2020;61(3):336–348.

48. Boer M, Stevens G, Finkenauer C, van den Eijnden R. Attention deficit hyperactivity disorder-symptoms, social media use intensity, and social media use problems in adolescents: investigating directionality. Child Dev. 2019;91(4):e853–865.

49. Trafford AM, Carr MJ, Ashcroft DM, et al. Temporal trends in eating disorder and self-harm incidence rates among adolescents and young adults in the UK in the 2 years since the onset of the COVID-19 pandemic: a population-based study. Lancet Child Adol Health. 2023;7(8):544–554.

50. Dixon D, Sharp CA, Hughes K, Hughes JC. Parental technoference and adolescents’ mental health and violent behaviour: a scoping review. BMC Public Health. 2023;23:2053.

