## Supplementary Material for "Excessive Internet Use among Finnish Adolescents – Prevalence in 2017–2023 and Associations with Well-being"

### Table S1. Distributions of the EIUS items among Finnish youth[a] in 2023, N = 149 298.

| EIUS item | Never % | Not very often % | Fairly often % | Very often % |
|---|---|---|---|---|
| I have tried unsuccessfully to spend less time online (relapse) | 43 962 (30.7) | 53 816 (37.6) | 32 850 (23.0) | 12 481 (8.7) |
| I have spent less time than I should have with family, friends, or doing homework because of the time I spent online (conflict) | 48 046 (33.7) | 50 826 (35.6) | 32 837 (23.0) | 10 976 (7.7) |
| I have caught myself surfing when I am not really interested (tolerance) | 40 082 (28.1) | 45 359 (31.8) | 42 267 (29.6) | 14 846 (10.4) |
| I have felt bothered when I cannot be online (withdrawal symptoms) | 68 949 (48.4) | 48 009 (33.7) | 18 506 (13.0) | 7041 (4.9) |
| I have gone without eating or sleeping because of the internet (salience) | 86 803 (61.0) | 36 685 (25.8) | 13 071 (9.2) | 5767 (4.1) |

EIUS = Excessive Internet Use Scale
a Including students in grades 8–11 (ages 13–20).

**Table S2. Associations of the EIUS symptoms with presence of EIU among Finnish youth[a] in 2023, N = 149 298.**

| EIUS symptom (item present fairly or very often) | No EIU<br>% | With EIU<br>% | Total<br>% |
|---|---|---|---|
| I have tried unsuccessfully to spend less time online (relapse) | 67.7 | 32.3 | 100.0 |
| I have spent less time than I should have with family, friends, or doing homework because of the time I spent online (conflict) | 63.8 | 36.2 | 100.0 |
| I have caught myself surfing when I am not really interested (tolerance) | 72.8 | 27.2 | 100.0 |
| I have felt bothered when I cannot be online (withdrawal symptoms) | 45.7 | 54.3 | 100.0 |
| I have gone without eating or sleeping because of the internet (salience) | 38.8 | 61.2 | 100.0 |

EIUS = Excessive Internet Use Scale
EIU = Excessive Internet Use (responding “fairly often” or “very often” to the conflict and at least three other items).
[a] Including students in grades 8–11 (ages 13–20).

**Table S3. Prevalence and 95% confidence intervals (CI) of EIU between 2017 and 2023 by gender and grade.**

| | 2017 (n = 133 731) | 2019 (n = 153 953) | 2021 (n = 159 459) | 2023 (n = 149 298) |
|---|---|---|---|---|
| **Subgroup/school level** | % (95% CI) | % (95% CI) | % (95% CI) | % (95% CI) |
| **Boys** | | | | |
| Grades 8 and 9 | 8.9 (8.6–9.2) | 8.6 (8.3–8.8) | 8.5 (8.2–8.7) | 9.1 (8.8–9.4) |
| Grades 10 and 11[a] | 6.2 (5.9–6.4) | 7.2 (7.0–7.5) | 7.2 (6.9–7.5) | 7.8 (7.5–8.1) |
| **Girls** | | | | |
| Grades 8 and 9 | 7.7 (7.5–8.0) | 9.6 (9.3–9.9) | 12.4 (12.1–12.8) | 14.0 (13.7–14.4) |
| Grades 10 and 11[a] | 5.8 (5.5–6.0) | 8.2 (7.9–8.5) | 10.5 (10.2–10.8) | 12.2 (11.8–12.5) |
| **Total** | | | | |
| Grades 8 and 9 | 8.3 (8.1–8.5) | 9.1 (8.9–9.3) | 10.6 (10.4–10.8) | 11.7 (11.5–11.9) |
| Grades 10 and 11[a] | 6.0 (5.8–6.2) | 7.8 (7.6–8.0) | 9.0 (8.8–9.2) | 10.2 (9.9–10.4) |

[a] Includes students in both academic high school and vocational education.
EIU = Excessive Internet Use, based on the 5-item EIU scale.

### Table S4. Prevalence and 95% confidence intervals (CI) of EIU between 2017 and 2023 by gender and grade. *Sensitivity analysis with complete cases*

| | 2017 (n = 115 792) | 2019 (n = 135 412) | 2021 (n = 141 801) | 2023 (n = 134 477) |
|---|---|---|---|---|
| **Subgroup/school level** | % (95% CI) | % (95% CI) | % (95% CI) | % (95% CI) |
| **Boys** | | | | |
| Grades 8 and 9 | 8.3 (8.0–8.6) | 7.8 (7.6–8.1) | 7.9 (7.7–8.2) | 8.5 (8.2–8.8) |
| Grades 10 and 11[a] | 5.7 (5.4–6.0) | 6.8 (6.5–7.0) | 6.8 (6.5–7.1) | 7.4 (7.1–7.7) |
| **Girls** | | | | |
| Grades 8 and 9 | 7.5 (7.2–7.8) | 9.5 (9.2–9.8) | 12.4 (12.0–12.7) | 13.9 (13.5–14.2) |
| Grades 10 and 11[a] | 5.7 (5.4–5.9) | 8.1 (7.8–8.4) | 10.5 (10.2–10.8) | 12.1 (11.8–12.5) |
| **Total** | | | | |
| Grades 8 and 9 | 7.9 (7.7–8.1) | 8.7 (8.5–8.9) | 10.3 (10.1–10.5) | 11.3 (11.1–11.6) |
| Grades 10 and 11[a] | 5.7 (5.5–5.9) | 7.5 (7.3–7.7) | 8.8 (8.6–9.0) | 10.0 (9.8–10.2) |

[a] Includes students in both academic high school and vocational education.
EIU = Excessive Internet Use, based on the 5-item EIU scale.

***Interpretation***

This is a sensitivity analysis where the results presented in Table S3 (see above) are reproduced using a dataset with no missing values allowed in any of the study variables used in analyses covering years from 2017 to 2023 (complete cases). Reductions in the number of cases when not allowing any missingness in these variables ranged from 14 821 (9.9%) in 2023 to 18 541 (12.0%) in 2019.

Differences between prevalence estimates using all available data (Table S3) and complete cases (Table S4) were negligible for the most. Largest changes were among boys from grades 8–9 and grades 10–11, where estimates reduced with 0.4–0.8 percentage points. The overall trend (differences between years) which was the main interest here, was not affected.

**Table S5. Odds ratios (OR) from adjusted logistic regression models for EIU as a predictor of mental and cognitive health outcomes among students in grades 8–11 in 2023. *Sensitivity analysis with complete cases.***

| | **Boys** | | **Girls** | |
|---|---|---|---|---|
| Outcome (dependent variable) | OR (95% CI)[b] | p | OR (95% CI)[b] | p |
| Sleep per weeknight <7 h | 1.76 (1.62–1.92) | < 0.001 | 2.10 (2.00–2.23) | < 0.001 |
| Depressive symptoms | 1.98 (1.82–2.16) | < 0.001 | 2.28 (2.18–2.39) | < 0.001 |
| Generalized anxiety symptoms | 2.42 (2.20–2.66) | < 0.001 | 2.20 (2.10–2.31) | < 0.001 |
| Social anxiety symptoms | 1.77 (1.64–1.90) | < 0.001 | 1.95 (1.86–2.04) | < 0.001 |
| Difficulties in tasks requiring | | | | |
| Reading | 1.95 (1.82–2.10) | < 0.001 | 1.96 (1.87–2.05) | < 0.001 |
| Writing | 1.88 (1.75–2.02) | < 0.001 | 1.95 (1.86–2.04) | < 0.001 |
| Mathematics | 1.85 (1.72–1.99) | < 0.001 | 1.99 (1.90–2.08) | < 0.001 |
| Difficulties concentrating | 2.35 (2.14–2.57) | < 0.001 | 2.16 (2.06–2.27) | < 0.001 |

[a] Adjusted for grade (8, 9, 10, and 11), living with both parents, family origin, parental education, and family financial situation, in total sample models also for gender.
[b] Separate model for each outcome; OR for EIU predicting the outcome.
EIU = Excessive Internet Use, based on the 5-item EIU scale.

***Interpretation***

This is a sensitivity analysis where the results presented in Table 4 of the main article for boys and girls are reproduced using a dataset with no missing values allowed in any of the study variables in 2023 (complete cases). In the analyses in Table 4 in the main article the number of cases ranged from 61 514 to 62 151 in boys and from 72 085 to 72 761 in girls depending on the outcome studied. In the complete case analysis presented in Table S5, there were 57 628 boys and 68 407 girls, thus number of missing cases due to missing values in the study variables ranged between 3886 and 4523 in boys and 3678 and 4354 in girls.

Differences between estimates using all available data (Table 4) and complete cases (Table S5) were negligible with largest differences in the ORs being of magnitude .02 (difficulties in concentrating in boys). This suggests that missingness due to missing values in the studied variables has only very little effect on the presented estimates.
